# Exploring the effects of a lack of pocketmoney onthe learning activities of undergraduate health sciences students in their clinical year

**DOI:** 10.1101/19009985

**Authors:** Marema Jebessa Kumsa, Bizuayehu Nigatu Lemu, Taklehaimanot Mezgebe Ketema

**Affiliations:** Department of Medical Radiology Technology, College of Health Sciences, School of Medicine, Addis Ababa University, Addis Ababa, Ethiopia; Department of Medical Radiology Technology, College of Health Sciences, School of Medicine, Addis Ababa University, Addis Abab, Ethiopia

**Keywords:** Undergraduate students, higher education, pocket money, learning, Ethiopia, health science, socioeconomic status, clinical placement

## Abstract

**Background:** In Ethiopia, there is little understanding of students’ university expenses and how the lack of pocket money affects undergraduate learning. This is particularly evident for undergraduate health sciences students that need additional money when they start their clinical placement.

**Objectives:** We aimed to explore the economic challenges that clinical year undergraduate health sciences students face with limited pocket money, as well as how students perceive these limited funds as affecting their learning activities and their ability to meet challenges.

**Methods:** This descriptive qualitative study was conducted at the Department of Medical Radiology Technology (MRT), College of Health Sciences (CHS) at Addis Ababa University (AAU). Ten participants were recruited through purposive and snowball methods, and semi-structured interviews were conducted between January 28, 2019 and February 1, 2019. The semi-structured questions explored participants’ experiences and perceptions regarding the challenges of a lack of pocket money and its impacts on their learning activities. Their reaction to financial challenges was also assessed.

**Results:** Four themes that are related to impact of lack of money on learning activities emerged from the interviews and they revolved around the students’ challenges in obtaining pocket money, their difficulty in affording daily needs, their self-management and their ability to have an adequate social life.

**Conclusion:** Based on our data, the underlying causes that lead students to face financial hardships can be solved by increasing public awareness of university expenses, clarifying the cost sharing system to the public, redesigning the cost sharing policy, and improving university services In addition, training students self-management and pocket money management, as well giving providing adequate transportation services, cash-money, and consultations for students during clinical placement.

## Introduction

The United Nations Educational, Scientific and Cultural Organization’s World Conference on Higher Education (1) states that “higher education needs to be a fundamental right for all,” regardless of a student’s economic status. However, students from low socioeconomic status (SES) backgrounds have lower educational aspirations, persistence rates, and educational achievements during college (2-5). Moreover, several previous studies suggest that during clinical placements; students face increased financial costs such as transportation, food expenses, clothing, and other material needs (6-8)

In Ethiopia, we currently lack an understanding of how insufficient discretionary funds for undergraduate health sciences students from low SES backgrounds impacts their learning activities within a clinical setting. Ethiopian Higher Education Proclamation and the Cost Sharing Regulations(9), cost-sharing is an open policy: provides food, dormitory and tution fee; however, it does not allow students to get discretionary money. Pocket money may be defined as the income that the student receives from a parent or guardian(10) and this definition used in this paper.

The aim of this study was to explore the impact of a lack of pocket money on undergraduate health sciences students’ learning activities during their years in the clinic. This research helped identify the challenges that undergraduate health sciences students faced due to the lack of pocket money and the perceived effects of their financial hardships on their learning activities.

## Methods

### Study design and setting

This study was a descriptive qualitative study that was used to explore the impacts of a lack of pocket money on undergraduate students’ clinical learning activities. Semi-structured interviews were used to gather data and the questions aimed to determine the students’ perceptions of the effects of their financial hardships on their learning activities. This study was conducted with third- and fourth-year students from the Department of MRT, College of Health Sciences, AAU in Ethiopia.

### Ethical considerations

Ethical clearance was obtained from AAU’s Department of Medical Education Ethical Review Committee. Informed consent was obtained from each study participant.

### Participant recruitment and data collection

We used purposive and snowball sampling techniques for participant recruitment. First, we invited those who have submitted letters to the department seeking financial support. Upon their arrival, they were briefed on the project and asked for self-declaration of their financial status. Then, snowball sampling was used in order to obtain other potential study participants. During all interviews, participants were asked to self-declare their financial status.

In the end, five participants from year four and seven participants from year three of their health sciences degree were interviewed. During transcribing the participants’ interviews, we found that two of the participants self-declared that they were in financial hardship but it disproved from what they respond and thus they were dropped from the data. Therefore, interviews from ten participants were transcribed verbatim, and translated. The participants’ descriptions are summarized in Table1.

**Table 1:**
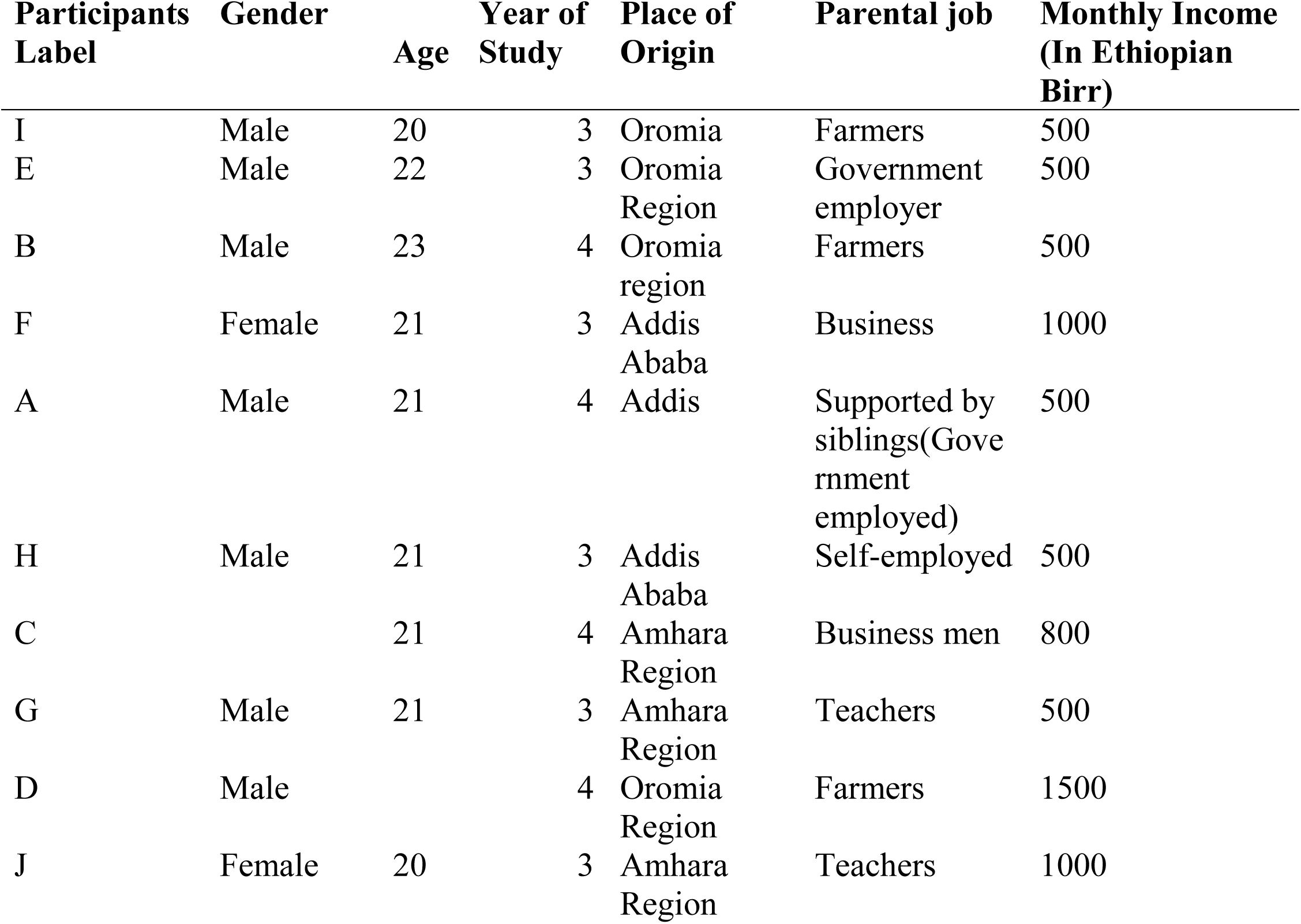
Participants’ characteristics.

### Interviews

The semi-structured interviews were designed using Pierre Bourdieu’s concepts of social capital (15). Two pilot interviews (not included in the analysis) were carried out to help refine the interview questions. An emphasis was placed on the lack of pocket money and its impacts on learning activities. The interviewer was selected from the Department of Nursing, and the interviews began on January 28, 2019 and were completed on February 1, 2019.The participants were interviewed in their language of choice (11 in Amharic and one in Afaan Oromo). Interview length ranged between 10 and 25 minutes. Participants were thanked for their participation and were given a notebook and 50 Ethiopian birr worth of mobile cards each.

### Data analysis

Researchers used a qualitative content analysis method (11-13), and thematically analyzed the interviews to explore the effects of low SES and a lack of discretionary money on health sciences students’ learning activities while in the clinic. The audio-recordings were transcribed verbatim and translated into English by the researchers. We held four meetings to transcribe and translate the data. The data were assessed by researchers at multiple levels. The three levels of data analyses (code, category, and theme) taken as appropriate for coding the data(13). After assessing the data, the codes, themes, and categories were formulated and were modified throughout the entire research process, with the full participation of all researchers. Table 2 contains a sample of the coded, categorized, and themed the data.

**Table 2:**
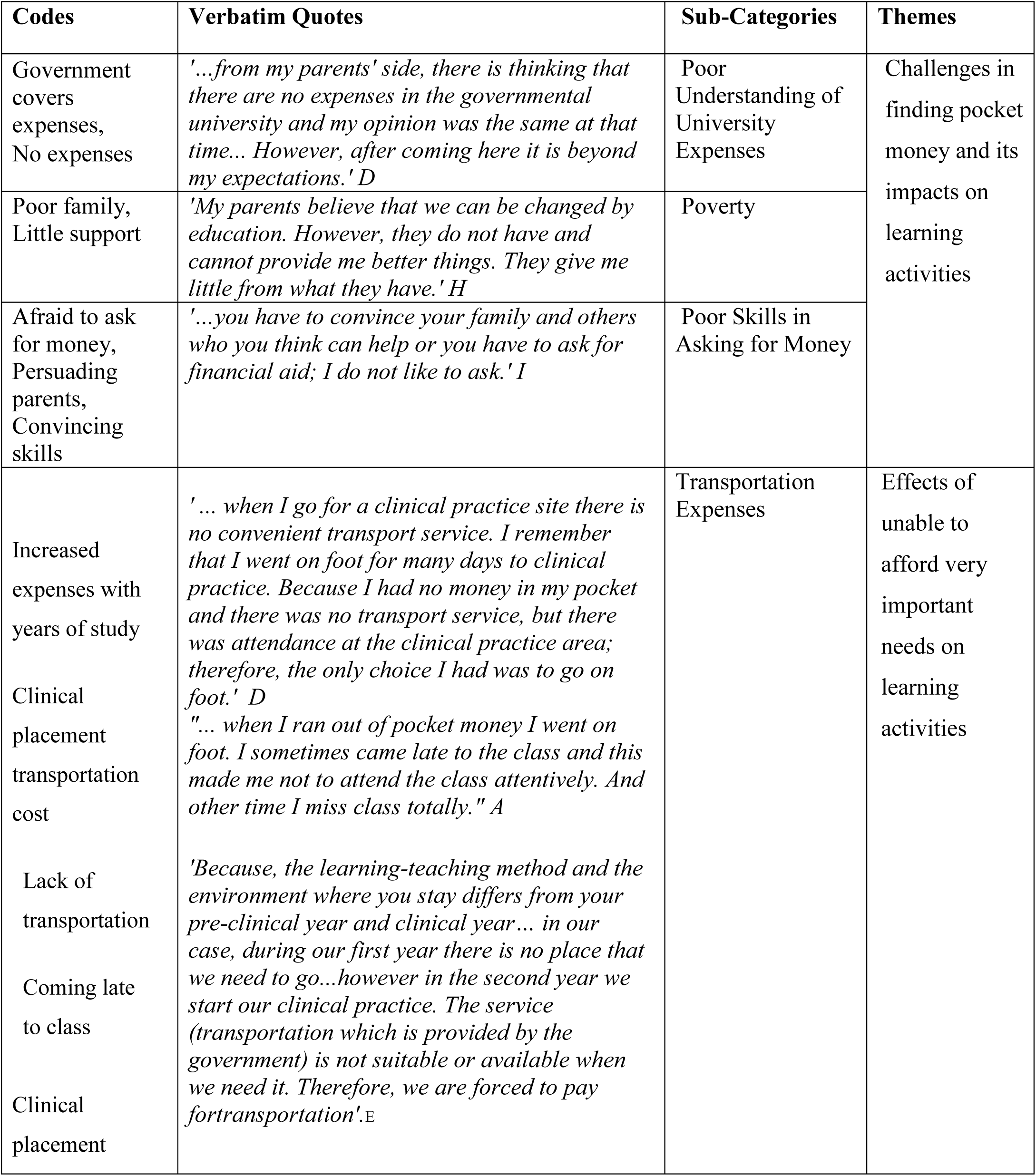
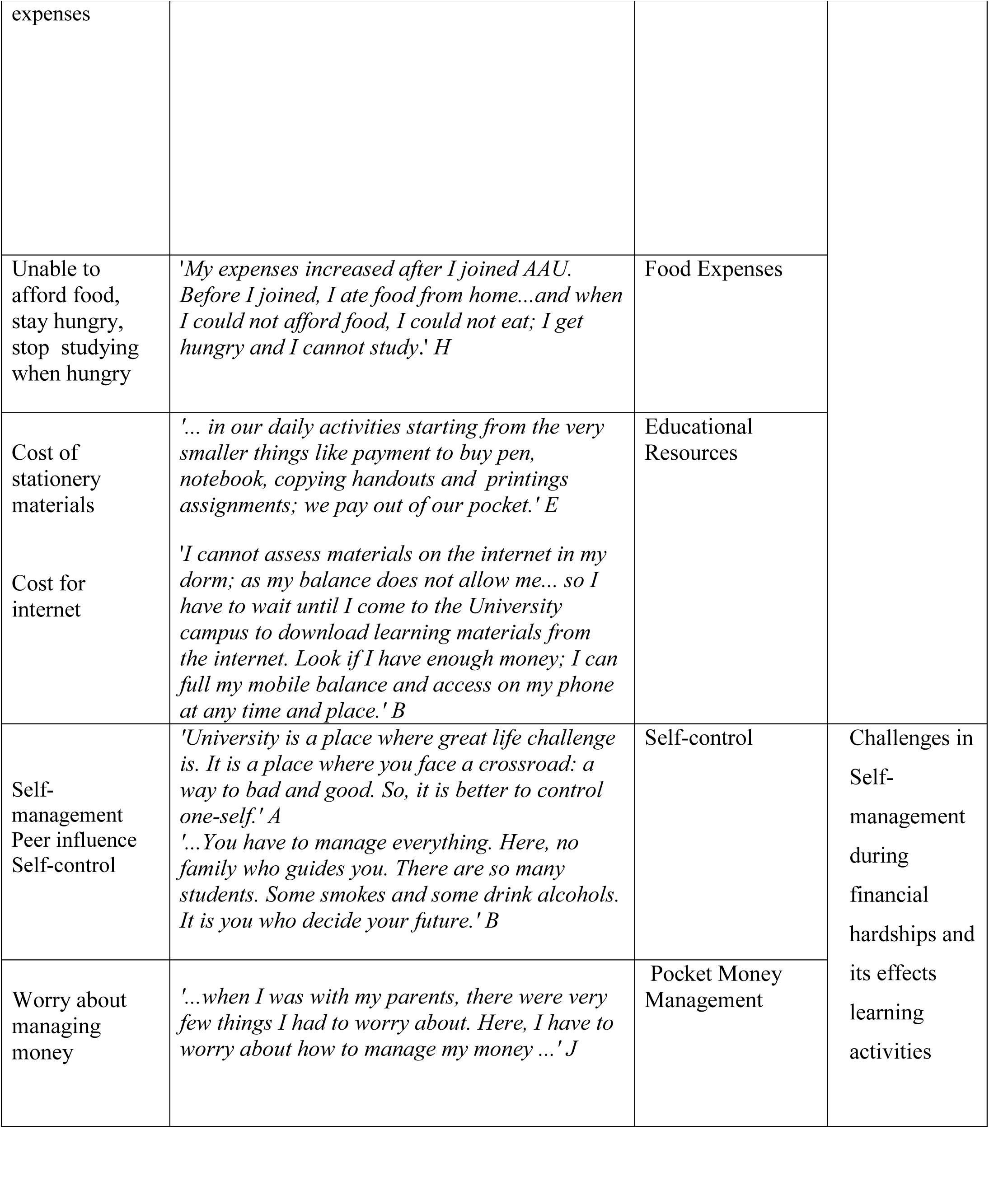
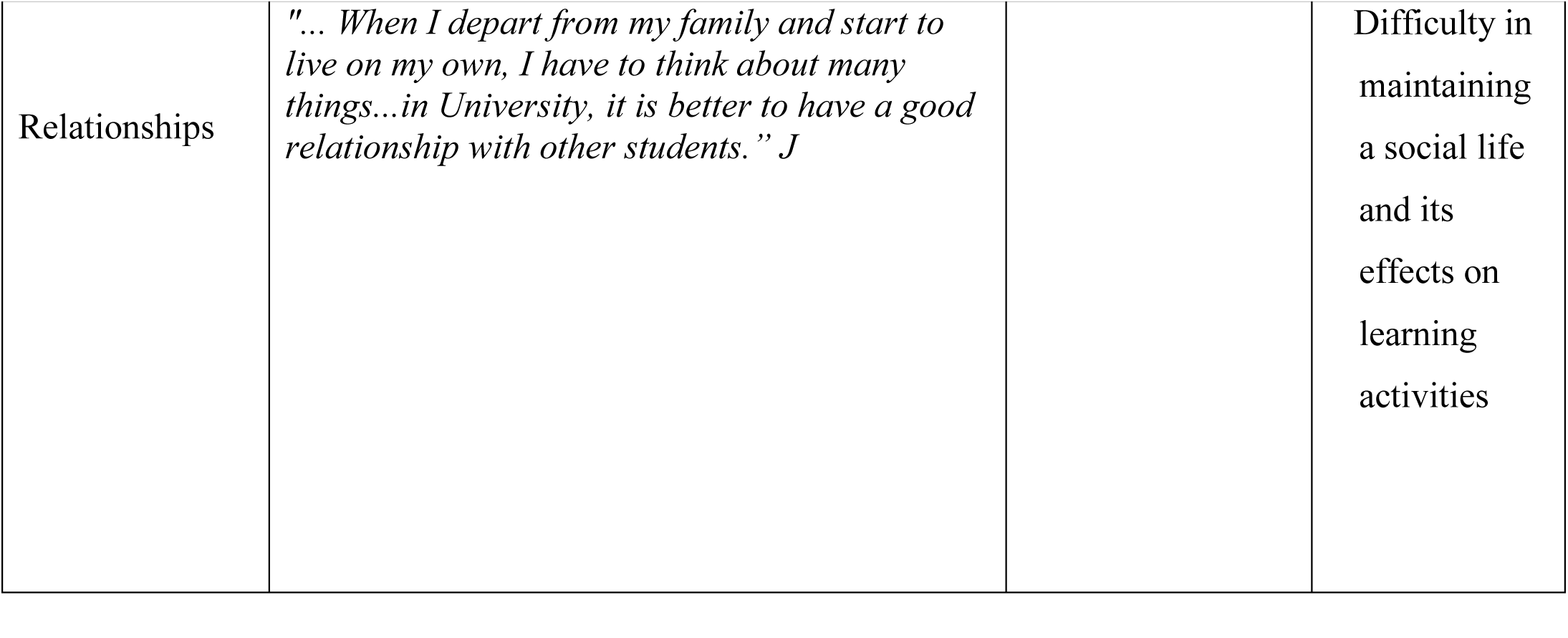
Summary of codes, categories, themes, and quotes.

Accordingly, four main themes explaining ways in which lack of pocket money affects learning activities were emerged. These are:

1. Challenges in finding pocket money and its impacts on learning activities
2. Effects of unable to afford very important needs on learning activities
3. Challenges in Self-management during financial hardships and its effects learning activities
4. Difficulty in maintaining a social life and its effects on learning activities

To explore the causes and challenges that students from low SES backgrounds face, their experiences and reactions to challenges, and the impacts of a lack of pocket money on learning activities, we applied Pierre Bourdieu’s concepts of habitus, cultural, and social capital (14) in analyzing the data.

### Theoretical approach: The Bourdieusian concept of habitus, cultural, and social capital

The Bourdieusian concept of habitus, cultural, and social capital was used to analyze the students’ experience of financial problems and its impacts on their learning activities. Bourdieu defines habitus as a property of actors that comprises a “structured and structuring structures”(14). Bourdieu defines habitus as one’s worldview, which is dictated by a person’s past and present circumstances, such as family upbringing and educational experiences.

Cultural capital refers to family-based cultural traits such as work habits, basic learning orientations, prevailing cultural norms, values, attitudes, and parenting styles and practices (15). This concept was developed by Pierre Bourdieu and Jean-Claude Passeron to analyze the impact of culture on the class system. This concept was developed by Pierre Bourdieu and Jean-Claude Passeron to assess the differences between student academic achievements (14).

Lastly, Bourdieu defines social capital as the ‘aggregate of actual or potential resources which are linked to possession of a durable network of more or less institutionalized relationships of mutual acquaintance and recognition’(14).With respect to social capital, proximity plays an important role in establishing a relationship.Proximity can be geographic in nature or it can be two persons’ closeness in blood, faith, profession, interest, study, culture, language, economy, or social proximity(16).

## Results

All participants discussed their perceived experiences of a lack of pocket money as undergraduate students and conceptualized its impact on their learning process. They also explained the means by which they overcame these problems in order to succeed in their studies. Participants’ responses were categorized into four themes, each of which was correlated to their effects on student learning activities:

1. Challenges in finding pocket money and its impacts on learning activities
2. Effects of unable to afford very important needs on learning activities
3. Challenges in Self-management during financial hardships and its effects learning activities
4. Difficulty in maintaining a social life and its effects on learning activities

### Theme 1: Challenges in finding pocket money and its impacts on learning activities

Nearly all of participants mentioned that getting pocket money for their various expenses in the university was a significant problem and identified a lack of awareness and absolute poverty as the main causes. Based on the data collected from the participants, we identified three categories that contribute to challenges in accessing enough discretionary money, and those categories are discussed below.

### Category 1: Poor understanding of university expenses

Several participants mentioned that most their parents have the idea that once their children join public universities, all their expenses are going to be paid by the government. The students identified this as being the most important factor that contributes to their financial troubles.

One of participant described the situation as:

> *‘…from my parents’ perspective, they thought that there were no expenses in a governmental university and my opinion was the same at that time. We talked about the government covering everything. However, after coming here (a governmental university) it was evident that this was not the case*.*’ D*.

And another participant said:

> *‘My parents believe that once you joined a university everything you need is provided by the government. And they assume that you only need money from them only for soaps and other smaller things*.*’ I*

These descriptions clearly show that there is a general misunderstanding of expenses for students at university among different families in the country.

### Category 2: Absolute parental Poverty

Some students explained that even though some of their parents know the expenses at university, their economic status does not allow them to cover those costs. This financial burden is a significant obstacle for those students in trying to get the most out of their education.

One of the participants described their case:

> *‘My parents believe that we can be changed by education. However, they do not have and cannot provide us with better things*.*’ H*

Absolute parental or related family members’ poverty directly affects student financial status and hence, students become unable to by pens, books, afford for printing of handouts, additional food and transportation to and from clinical practice sites. This has negative effects on their daily learning activities.

### Category 3: Poor skills in asking for money

Participants explained that considering their community’s understanding of university expenses, they felt that they must develop the ability to persuade parents and family members in order to get the pocket money. Unfortunately, several participants did not ask parents or family members for money because of the lack of cultural acceptance for that sort of behaviour.

One of participant described their experience as:

> *‘…you have to convince your family and others which we lack due to …the other is you have asked for financial aid…there is no well established financial aid*.*’ I*

Another participant also explained it as:

> *‘My parents do not know about handouts, assignments, and that I went to clinical placements using transportation…they do not know and have no experience with higher education*.*’ B*

To summarize many participants asking for pocket money is real problem and they mentioned that this has a significant contribution in making them not afford what they need. Thus, this has a huge impact on their learning activities.

### Theme 2: Effects of unable to afford very important needs on learning

Several study participants indicated that the lack of pocket money had a tremendous impact on their daily lives, particularly affecting their ability to accomplish their learning goals. Most participants stated that the small amount of pocket money that they had almost went to fulfilling basic needs. Based on the data, Theme 2 was divided into three categories and each of them are discussed below separately.

### Category 1: Transportation expenses

Many participants mentioned that most of their pocket money is spent on transportation. In addition to transportation between two campuses and from home to the university, these students must also absorb the cost of transportation to their clinical practice away from their campus. Some of the participants even mentioned that they were forced to go on foot to their clinic placement as there was no access to transportation from the university and they had run out of pocket money. This greatly influenced their ability to acquire the necessary clinical skills.

One participant said the following:

> *‘…when I received a clinical placement, I found that there was no convenient transportation service. I remember that I had to go on foot many times to my clinical practice because I had no money in my pocket…but attendance was taken at the clinical practice area*.*’ D*

Another participant said:

> *‘…when I ran out of pocket money I went on foot. Sometimes I arrived late to class and this greatly affected my focus in class. Another time I missed class altogether*.*’ A*

### Category 2: Food expenses

Food is one of the biggest expenses for university students. Several participants described different situations: unfavourable timing of food service from the university cafeteria, unfavourable food of the university cafeteria, during time of illness, during clinical placement and during night times. They explained that they buy food out of university campus when they had money and stay hungry when they run out of pocket money.

One of participants mentioned:

> ‘*My expenses increased after I joined AAU. Before I joined here, I ate food from home…and when I could not afford food here at the university, I stayed hungry and had a hard time studying*.’ *H*

Another participant said:

> *‘…for example, I use the university cafeteria. However, since the food is not very good, I sometimes eat outside the campus*.*’ C*

As described by several participants, a lack of pocket money forced them to go without food, which makes it very difficult to study or attend classes. Hence, a lack of pocket money can greatly influence a student’s ability to effectively get an education.

### Category 3: Educational resources

Educational resources can also be another significant drain on a student’s discretionary money. The educational resources on which participants spent their pocket money included necessities like stationery materials and internet access. These are further detailed below:

### Subcategory 1: Stationery materials

Participants indicated that they spent money on buying pens, notebooks, copying and printing handouts, and printing assignments almost daily.

One of the participants described student spending on stationary materials as the following:

> *‘…on a daily basis, we pay out of pocket for things such as pens, notebooks, copying handouts, and printing assignments*.*’ E*

Several participants indicated that their inability to afford educational materials affected their overall learning activities.

### Subcategory 2: Access to the internet

Many of the participants specified that the campus where they live does not provide internet access and thus they needed to either use mobile data or go to an internet café, both of which require money.

One participant said:

> ‘*I cannot assess materials on the internet in my dorm… so I have to wait until I come to another university campus to use the internet. Look, if I had enough money, I would just use my mobile phone’s internet access to get on the internet at any time and place*.*” B*

Another participant said:

> *‘…at our campus there is no good internet connectivity…if you want to use the internet you will need to pay for it ‘ E*

Here, the participants identified internet access as an important part of their learning process. Unfortunately, they do not have access to free internet services and are generally unable to pay for it out of pocket.

### Theme 3: Self-management

Several participants mentioned that leaving their family and living at the university was the most challenging aspect of getting their education. Most of them discussed it as it relates to challenges of self-control and pocket money management. These aspects are further explored below.

### Category 1: Self-control

University is a place where students start to exercise self-control. Most of the participants indicated that when someone does not have enough pocket money and has poor self-control it is very difficult to be successful at university. In addition, they mentioned that self-control helps reduce the stress that develops due to a lack of pocket money and therefore improves proper learning.

One participant mentioned:

> *“University is a place where a person confronts some of life’s greatest challenges. It is where one faces a crossroad: a way to bad and good. So, it is better to control oneself eventhough you struggle financially” A*

Another participant said:

> *“You have to manage everything. Here, there is no family around who to guide you. There are so many students and some smoke and some drink alcohol. You have to decide your future for yourself*.*” B*

Many of the participants focused on self-management and its importance in being successful at university. They believed that even though many students have financial problems it is possible to be successful at university if self-management is practiced effectively. Interestingly, most of the participants mentioned that they have developed good self-management.

### Category 2: Pocket money management

The effective use of the small amount of discretionary money that the students have while in university is very important, as was discussed by the participants.

One of participant said:

> *‘…One has to use the small amount of pocket money received sparingly. ‘ I*

Another participant mentioned:

> *‘In general, using the money provided to you effectively is very difficult, as is getting my education*.*’ B*

### Theme 4: Difficulty in maintaining a social life and its effects on learning activities

Participants believed that success in pursuing a higher education is highly dependent on having a reliable network of people around the student. They stated that their lack of pocket money affected their ability to make connections with peers and noted that the lack of a social life affected their learning activities.

One interviewee expressed:

> *“… When I left my family and started to live on my own, I had to think about many things. in University, it is better to have a good relationship with other students has it has many benefits*.*” J*

## Discussion

Various factors contribute to students’ financial challenges at university. Here, we investigated how the lack of pocket money influences students learning by discussing the topic with several health sciences students at AAU. More specifically, we used Bourdieu’s concepts of habitus, cultural, and social capital to explore what contributes to the financial pressures experienced by undergraduate health sciences students during their time in clinic, and the impact of these challenges on their daily learning activities.

This study showed that the familial habitus of the participants’ affected their life at university. Participants described that many of their challenges based from the significant changes that accompanied moving away from their home and into a university environment. For instance, they went from having their lives mainly managed by their parents and families to self-management. Moreover, they went from living with students with similar lifestyles to living with students of different backgrounds. With all these changes, the lack of pocket money added another stress to their university life.

Several participants mentioned that the absence of a government policy that entitles students to get financial support during their undergraduate studies increased their financial hardship, which is similar to the findings of a study done in the United Kingdom(8). These financial stresses can be buoyed by a positive social structure. For instance, a previous study demonstrated that students with low SES and good self-management benefit from gaining social capital(15). Findings from our study support this idea and indicate that there does indeed appear to be a link between social bonding and achieving relief from financial crisis. This was observed by several participants that described the benefits of developing a culture of borrowing from friends.

This study found that students perceived self-management as the most important thing to combat the financial pressures encountered at university. Bourdieu’s concept of habitus (14), describing the way of acting, feeling and being, is related to the participants idea of self-control and pocket money management.

Similar to study done in Australia(17), most participants in this study had expected financial hardship to be a substantial concern at the beginning of their degree, however many of them experienced acute financial hardship during clinical placements. We found that students, who relocated for educational purposes, including placements, were economically disadvantaged and experienced financial stress (6). Furthermore, this study indicated that financial stress increased student stress overall, which was also observed in Greece(18)and in another location in Ethiopia(19).Of course, this increased stress negatively impacted student learning, as it is known to do. Several Participants expressed that even though their families misunderstood the expenses associated with a higher education, they overcame the challenges and worked toward attaining their educational goals(20).

Unlike the studies presented by Bennett (21) and Trombitas (22), this study does not support the idea that financial pressure causes students to drop-out or quit their education. Instead of dropping out the participants reported different methods of overcoming their acute financial problems, including borrowing from friends and developing strong self-management skills (i.e., prioritizing their basic and urgent needs and compromising other non-essentials). That said, further research is necessary to determine the relationship between financial hardship for Ethiopian health sciences undergraduate students and dropout rates.

### Strengths and Limitations

The main limitation to this study is that it included students from one department and from one institution. However, with this limitation, the current study contributes to a broader overview of the effects of financial hardship on undergraduate success and provides a better understanding of its causes.

## Conclusion

This study revealed that student financial hardships at AAU occurred mainly due to the lack of understanding about university expenses from families, absolute poverty, and poor university administrative services. This study also indicated that the cost-sharing system is not helpful in supporting financially disadvantaged students. Moreover, the lack of student pocket money had a negative impact on students’ theoretical learning activities and clinical skill acquisition. These findings indicate that the department, the school, the college, and the university need to find adequate solutions to these problems. This study also recommends that the Ethiopian government, specifically the Ministry of Science and Higher Education, should revise the Ethiopian cost-sharing system.

## Data Availability

All relevant data are within the manuscript and its Supporting Information files.

## Abbreviations

AAU: **Addis Ababa University**
CHS: College of Health Sciences
MRT: Medical Radiologic Technology
SES: socioeconomic status

